## Appendix for "Adoption and continued use of mobile contact tracing technology: Multilevel explanations from a three-wave panel survey and linked data"

**Figure A1**: Missing data patterns across attitudinal predictors + adoption (DV, wave 2)

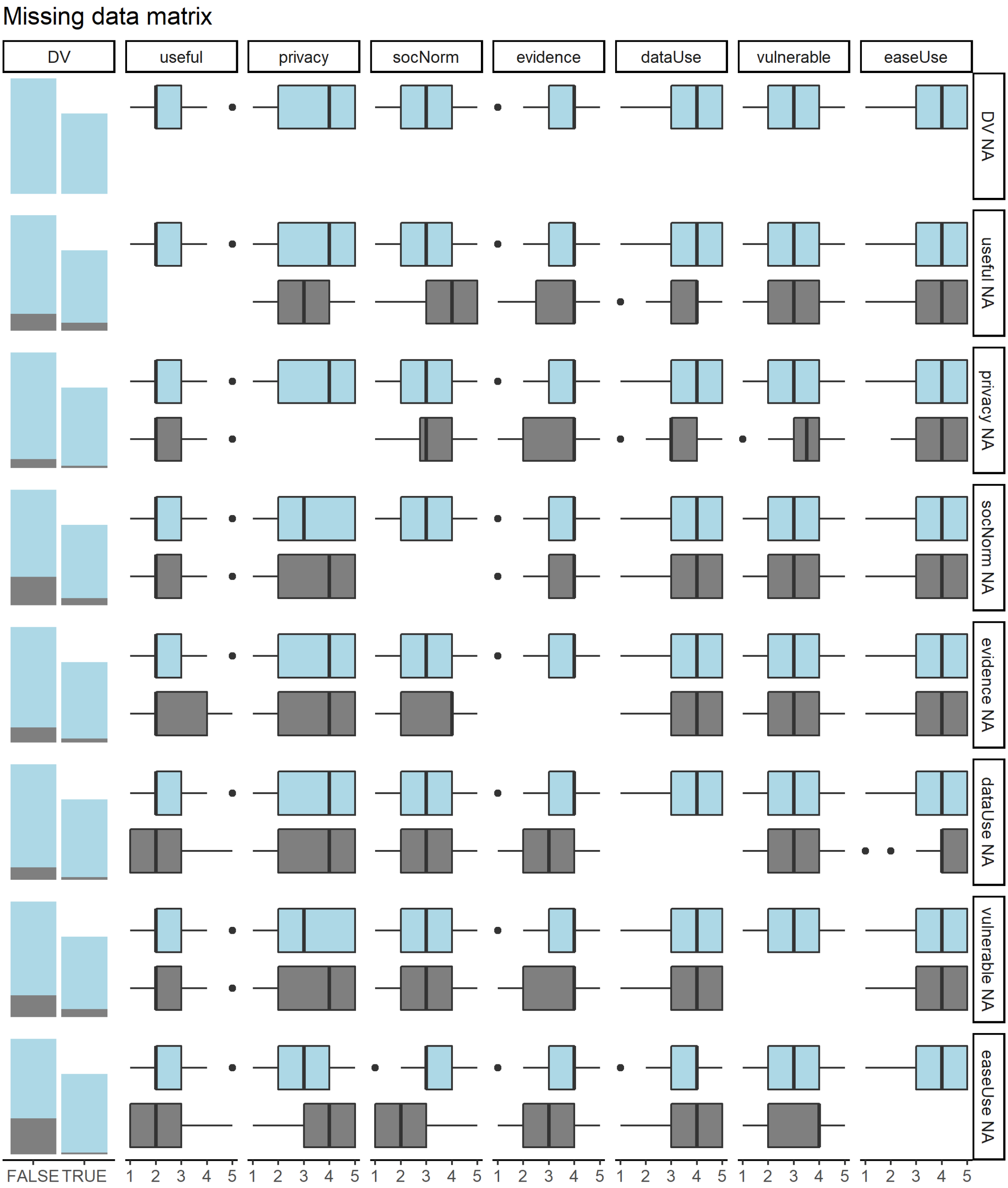


*Notes: attitudinal predictors more likely to be missing for non-adopters, particularly "ease of use"; some but not all attitudes are related to each other in terms of missingness, for example those with missing data on ease of use were more concerned about privacy.*

**Table A1:** Logistic regression results pooled across five multiply imputed*** datasets

| Model | Term | Log odds | OR | SE | p | RIV* | Lambda** |
|---|---|---|---|---|---|---|---|
| | Intercept | -0.82 | 0.44 | 0.23 | 0.00 | 0.02 | 0.02 |
| | Age | -0.18 | 0.84 | 0.06 | 0.00 | 0.04 | 0.04 |
| | Women | 0.01 | 1.01 | 0.10 | 0.89 | 0.01 | 0.01 |
| | Education | 0.00 | 1.00 | 0.11 | 0.97 | 0.06 | 0.06 |
| **Initial adoption** | Ethnic minorities | -0.03 | 0.97 | 0.19 | 0.87 | 0.04 | 0.04 |
| N = 2,500 | Perceived usefulness | -0.03 | 0.97 | 0.06 | 0.61 | 0.42 | 0.30 |
| N (districts) = 1,282 | Privacy concern | -0.81 | 0.44 | 0.07 | 0.00 | 0.05 | 0.05 |
| SD (districts) = 0.34 | Social norms | 0.60 | 1.82 | 0.06 | 0.00 | 0.56 | 0.36 |
| m = 5*** | Transparent evidence | 0.13 | 1.13 | 0.06 | 0.05 | 0.14 | 0.12 |
| | More info on data usage | 0.12 | 1.12 | 0.07 | 0.13 | 0.19 | 0.16 |
| AIC = 2596.07 | Evidence on vulnerable groups | 0.01 | 1.01 | 0.06 | 0.89 | 0.15 | 0.13 |
| BIC = 2712.55 | Ease of use | 0.61 | 1.84 | 0.06 | 0.00 | 0.14 | 0.12 |
| logLik = -1278.03 | Trust in UK Gov't | -0.04 | 0.96 | 0.05 | 0.43 | 0.04 | 0.04 |
| df.resid = 2,480 | Mobility | -0.20 | 0.82 | 0.11 | 0.08 | 0.04 | 0.04 |
| | Compliance | 0.29 | 1.33 | 0.12 | 0.02 | 0.04 | 0.04 |
| | Tier 2 (district) | -0.19 | 0.83 | 0.11 | 0.10 | 0.01 | 0.01 |
| | Tier 3 (district) | -0.20 | 0.82 | 0.16 | 0.22 | 0.03 | 0.03 |
| | Cases (district) | 0.03 | 1.03 | 0.05 | 0.60 | 0.07 | 0.07 |
| | Urban (district) | 0.33 | 1.39 | 0.18 | 0.08 | 0.01 | 0.01 |
| | Intercept | 1.67 | 5.29 | 0.28 | 0.00 | 0.01 | 0.01 |
| | Age | 0.12 | 1.13 | 0.11 | 0.28 | 0.01 | 0.01 |
| | Women | 0.17 | 1.19 | 0.21 | 0.41 | 0.01 | 0.01 |
| | Education | 0.28 | 1.32 | 0.21 | 0.19 | 0.01 | 0.01 |
| **Continued use** | Ethnic minorities | -0.28 | 0.76 | 0.35 | 0.43 | 0.00 | 0.00 |
| N = 1,026 | Perceived usefulness | 0.16 | 1.18 | 0.11 | 0.16 | 0.10 | 0.09 |
| m = 5*** | Privacy concern | -0.26 | 0.77 | 0.13 | 0.05 | 0.03 | 0.03 |
| | Social norms | 0.27 | 1.31 | 0.13 | 0.06 | 0.27 | 0.21 |
| AIC = 742.12 | Transparent evidence | -0.25 | 0.78 | 0.14 | 0.08 | 0.09 | 0.08 |
| BIC = 816.12 | More info on data usage | 0.00 | 1.00 | 0.14 | 1.00 | 0.10 | 0.09 |
| logLik = -356.06 | Evidence on vulnerable groups | -0.03 | 0.97 | 0.12 | 0.84 | 0.12 | 0.11 |
| df.resid = 1,011 | Ease of use | 0.20 | 1.22 | 0.13 | 0.13 | 0.03 | 0.03 |
| | Trust in UK Gov't | -0.04 | 0.96 | 0.11 | 0.69 | 0.01 | 0.01 |
| | Mobility | 0.06 | 1.06 | 0.22 | 0.78 | 0.00 | 0.00 |
| | Compliance | -0.16 | 0.85 | 0.22 | 0.45 | 0.01 | 0.01 |
| | Intercept | -3.31 | 0.04 | 0.32 | 0.00 | 0.02 | 0.01 |
| | Age | -0.49 | 0.62 | 0.12 | 0.00 | 0.02 | 0.01 |
| | Women | 0.29 | 1.34 | 0.22 | 0.19 | 0.00 | 0.00 |
| | Education | 0.52 | 1.68 | 0.23 | 0.02 | 0.01 | 0.01 |
| **New adoption** | Ethnic minorities | -0.22 | 0.80 | 0.38 | 0.56 | 0.01 | 0.01 |
| N = 1,474 | Perceived usefulness | 0.04 | 1.04 | 0.12 | 0.77 | 0.07 | 0.07 |
| m = 5*** | Privacy concern | -0.37 | 0.69 | 0.15 | 0.02 | 0.02 | 0.02 |
| | Social norms | 0.34 | 1.40 | 0.13 | 0.02 | 0.16 | 0.14 |
| AIC = 688.88 | Transparent evidence | 0.18 | 1.20 | 0.14 | 0.19 | 0.08 | 0.08 |
| BIC = 768.32 | More info on data usage | 0.06 | 1.07 | 0.15 | 0.69 | 0.12 | 0.11 |
| logLik = -329.44 | Evidence on vulnerable groups | 0.08 | 1.08 | 0.13 | 0.55 | 0.05 | 0.05 |
| df.resid = 1,459 | Ease of use | -0.04 | 0.96 | 0.13 | 0.77 | 0.31 | 0.24 |
| | Trust in UK Gov't | 0.34 | 1.40 | 0.12 | 0.01 | 0.01 | 0.01 |
| | Mobility | 0.29 | 1.34 | 0.25 | 0.25 | 0.00 | 0.00 |
| | Compliance | 0.11 | 1.12 | 0.23 | 0.63 | 0.00 | 0.00 |

* Relative increase in variance due to missingness
** Proportion of total variance due to missingness
*** Number of multiple imputations generated with Gibbs sampling (predictive means) implemented in `mice` (Van Buuren and Groothuis-Oudshoorn, 2011).
