## Supplementary material for "Adoption and continued use of mobile contact tracing technology: Multilevel explanations from a three-wave panel survey and linked data": SQUIRE

### Reporting checklist for quality improvement in health care.

Based on the SQUIRE guidelines.

| | | Reporting Item | Page Number |
|---|---|---|---|
| **Title** | | | |
| | #1 | Indicate that the manuscript concerns an initiative to improve healthcare (broadly defined to include the quality, safety, effectiveness, patientcenteredness, timeliness, cost, efficiency, and equity of healthcare) | 2,9 |
| **Abstract** | | | |
| | #02a | Provide adequate information to aid in searching and indexing | 1 |
| | #02b | Summarize all key information from various sections of the text using the abstract format of the intended publication or a structured summary such as: background, local problem, methods, interventions, results, conclusions | 1 |
| **Introduction** | | | |
| Problem description | #3 | Nature and significance of the local problem | 2,3 |
| Available knowledge | #4 | Summary of what is currently known about the problem, including relevant previous studies | 2,3 |
| Rationale | #5 | Informal or formal frameworks, models, concepts, and / or theories used to explain the problem, any reasons or assumptions that were used to develop the intervention(s), and reasons why the intervention(s) was expected to work | 3 |
| Specific aims | #6 | Purpose of the project and of this report | 3 |
| **Methods** | | | |
| Context | #7 | Contextual elements considered important at the outset of introducing the intervention(s) | 3,4 |

| | | | |
|---|---|---|---|
| Intervention(s) | #08a | Description of the intervention(s) in sufficient detail that others could reproduce it | NA |
| Intervention(s) | #08b | Specifics of the team involved in the work | NA |
| Study of the Intervention(s) | #09a | Approach chosen for assessing the impact of the intervention(s) | NA |
| Study of the Intervention(s) | #09b | Approach used to establish whether the observed outcomes were due to the intervention(s) | NA |
| Measures | #10a | Measures chosen for studying processes and outcomes of the intervention(s), including rationale for choosing them, their operational definitions, and their validity and reliability | 4 to 6 |
| Measures | #10b | Description of the approach to the ongoing assessment of contextual elements that contributed to the success, failure, efficiency, and cost | 4 to 6 |
| Measures | #10c | Methods employed for assessing completeness and accuracy of data | 6 |
| Analysis | #11a | Qualitative and quantitative methods used to draw inferences from the data | 6 |
| Analysis | #11b | Methods for understanding variation within the data, including the effects of time as a variable | 6 |
| Ethical considerations | #12 | Ethical aspects of implementing and studying the intervention(s) and how they were addressed, including, but not limited to, formal ethics review and potential conflict(s) of interest | 2 |
| **Results** | | | |
| | #13a | Initial steps of the intervention(s) and their evolution over time (e.g., time-line diagram, flow chart, or table), including modifications made to the intervention during the project | NA |
| | #13b | Details of the process measures and outcome | 6,7 |
| | #13c | Contextual elements that interacted with the intervention(s) | 7 |
| | #13d | Observed associations between outcomes, interventions, and relevant contextual elements | 7,8,A2 |
| | #13e | Unintended consequences such as unexpected benefits, | 8,9 |

| | | | |
|---|---|---|---|
| | | problems, failures, or costs associated with the intervention(s). | |
| | #13f | Details about missing data | A1 |
| **Discussion** | | | |
| Summary | #14a | Key findings, including relevance to the rationale and specific aims | 9 |
| Summary | #14b | Particular strengths of the project | 1,2,9 |
| Interpretation | #15a | Nature of the association between the intervention(s) and the outcomes | 9 |
| Interpretation | #15b | Comparison of results with findings from other publications | 9 |
| Interpretation | #15c | Impact of the project on people and systems | 9 |
| Interpretation | #15d | Reasons for any differences between observed and anticipated outcomes, including the influence of context | 9 |
| Interpretation | #15e | Costs and strategic trade-offs, including opportunity costs | NA |
| Limitations | #16a | Limits to the generalizability of the work | 1,2,9 |
| Limitations | #16b | Factors that might have limited internal validity such as confounding, bias, or imprecision in the design, methods, measurement, or analysis | 1,2,A1 |
| Limitations | #16c | Efforts made to minimize and adjust for limitations | 6,A1,A2 |
| Conclusion | #17a | Usefulness of the work | 9 |
| Conclusion | #17b | Sustainability | 9 |
| Conclusion | #17c | Potential for spread to other contexts | 6 |
| Conclusion | #17d | Implications for practice and for further study in the field | 9 |
| Conclusion | #17e | Suggested next steps | 2,9 |
| **Other information** | | | |
| Funding | #18 | Sources of funding that supported this work. Role, if any, of the funding organization in the design, implementation, interpretation, and reporting | 2 |

The SQUIRE 2.0 checklist is distributed under the terms of the Creative Commons Attribution License CC BY-NC 4.0. This checklist was completed on 10. May 2021 using https://www.goodreports.org/, a tool made by the EQUATOR Network in collaboration with Penelope.ai
